# Association of Mid-Infrared Optoacoustic Sensing with Glucose Dynamics in Humans

**DOI:** 10.64898/2026.11.24.26368846

**Authors:** Alexander Prebeck, Maximilian Gotsch, Francesca Gasparin, Ludwig Englert, Uli Stahl, Vasilis Ntziachristos

**Author notes:** Corresponding Author: V.N.

## Abstract

Despite transformative advances in continuous glucose monitoring, current continuous glucose monitoring (CGM) still relies on transcutaneous needle-based sensors that measure glucose in the interstitial fluid, which lags behind blood glucose. Mid-infrared optoacoustic sensing (MiROS) has recently demonstrated a sensitivity to glucose changes of a few tens of mg/dL in mice using non-invasive measurements. However, the relation of MiROS to human measurements remains unknown. We conducted a MiROS pilot study in 30 healthy participants undergoing oral glucose tolerance tests and investigated how optoacoustic signals reflect tissue optical-property variations associated with changes in blood glucose concentration. We defined the Optoacoustic Mid-infrared Signal (OMS) as a readout of glucose-induced alterations in tissue optical properties and compared it with reference blood glucose measurements. The OMS corresponded to a mean absolute error (MAE) of 11.9 mg/dL and a mean absolute percentage error (MAPE) of 10% relative to the reference blood analysis. More than 99.9% of paired OMS values were located in zones A and B of the consensus error grid. The study further showed that OMS compared favourably with concurrently acquired CGM measurements, which showed an MAE of 15.7 mg/dL and a MAPE of 12% and exhibited an average time lag of 10.5 min relative to blood glucose. No comparable lag was observed for the OMS. These results support MiROS as a promising framework pointing to a sensor with potential for non-invasive glucose sensing in humans, in the future, and a potential step toward less burdensome glucose monitoring for diabetes management.

## Introduction

Continuous glucose monitoring (CGM) has transformed glucose management by enabling near-continuous measurements throughout daily life. However, current CGMs measure glucose indirectly in interstitial fluid (ISF) rather than directly in blood, introducing physiological and measurement constraints that can limit accuracy, particularly during rapid glucose changes^1–4^. Despite substantial technological advances, *in vivo* sensing without needles remains an unsolved challenge and the holy grail of glucose detection. Fully non-invasive detection could improve comfort over needle-based approaches and potentially broaden adoption among both the more than 529 million people living with diabetes worldwide^5^, and possibly non-diabetic populations including high-risk and health-conscious individuals.

Recent progress with mid-infrared optoacoustic sensing (MiROS) has opened the possibility to eliminate skin penetration, while detecting glucose from tissue layers rich in blood. However, while MiROS’ performance was validated in mice^6^, its capabilities in humans remain unknown. Here, we report the first results from MiROS measurements performed on a cohort of 30 healthy participants, during an oral glucose tolerance test (OGTT). In a retrospective data analysis, we investigated in particular the relationship between temporal changes in the Optoacoustic Mid-infrared Signals (OMS) and corresponding changes of reference glucose concentrations that were determined on capillary blood samples, acquired in tandem with the MiROS measurements. We further compared the OMS with concurrent readings from a commercially available continuous glucose monitoring system and used exploratory regression analyses to identify participant characteristics and glucose dynamics that affect OMS performance.

We show that, in this cohort, the OMS followed the temporal evolution of the glucose concentration values determined by capillary blood analysis with higher accuracy than the concurrently acquired CGM measurements. These findings corroborate our previous observations in mice, in which MiROS signals closely followed the temporal dynamics of invasive reference blood glucose measurements without exhibiting the delays associated with the interstitial compartment. No statistically significant associations were detected between OMS accuracy and participant characteristics, including age, skin color, and body mass index (BMI).

This study provides initial evidence that glucose-associated tissue changes accurately map onto MiROS measurements obtained non-invasively from humans. We discuss possible implementations that can further increase the accuracy of the measurement in future system iterations.

## Results

### Study and Participant Distribution

To evaluate the relationship between the OMS and blood glucose in humans, we implemented a MiROS measurement system for interrogation of the earlobe (Fig. 1a). Pulsed mid-infrared irradiation on one side of the earlobe induced ultrasound waves through the optoacoustic effect. The ultrasound waves were detected on the opposite side of the earlobe, by a confocally aligned ultrasound transducer after propagation through the tissue (Fig. 1b).

**Figure 1:**
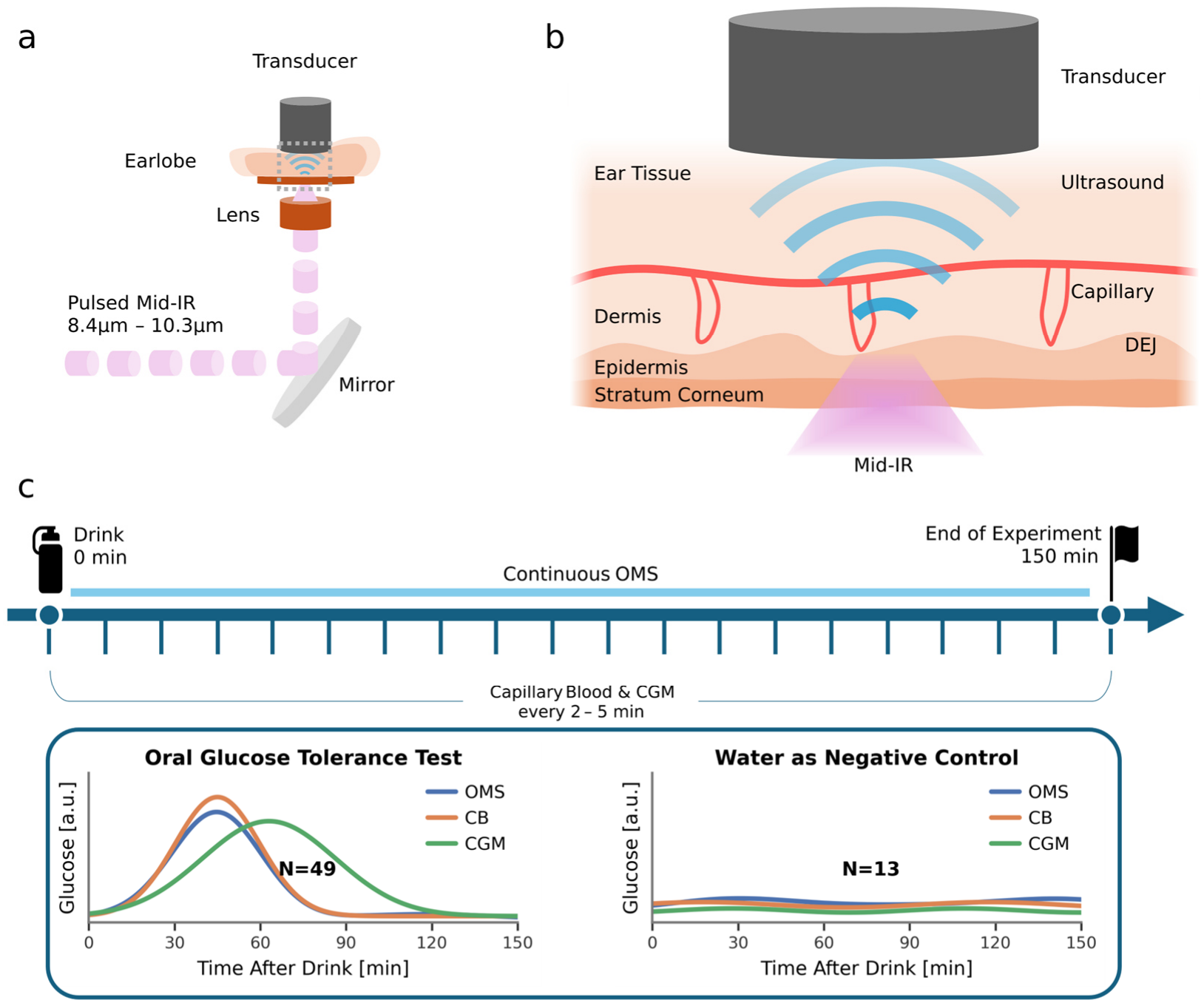
System and study overview. (a) Schematic of the mid-infrared optoacoustic sensing (MiROS) system used for optoacoustic mid-infrared signal (OMS) measurements on the earlobe. (b) Schematic of the geometrical arrangement of the OMS measurement in transillumination mode of the earlobe. (c) Study protocol showing a schematic overview of the measurement sessions, including ingestion of either glucose solution during an oral glucose tolerance test or plain water as a negative control. Mid-IR: mid-infrared, DEJ: epidermal-dermal junction, CGM: continuous glucose monitor, CB: capillary blood.

To assess the relation of the OMS recorded to the underlying glucose concentration, a cohort of 30 healthy participants underwent at least two measurement sessions (Fig. 1c). All participants ingested a glucose solution during the first session, whereas approximately one third received pure water in the second session. On each of the 62 scanning sessions, MiROS measurements were performed in parallel with reference measurements obtained by CGM and invasive capillary blood sampling. Each MiROS measurement lasted approximately one minute, whereby the capillary blood samples were obtained every two to five minutes. The cohort was approximately balanced by sex and heterogeneous with respect to age, skin tone, and BMI, enabling evaluation of MiROS performance across diverse physiological conditions (Fig. 2, Table 1).

**Table 1.** Descriptive statistics of participant characteristics in the study cohort.

| Participant Characteristic | Mean | Standard Deviation | **Minimum** | **Maximum** |
|---|---|---|---|---|
| Age [years] | 32.2 | 8.96 | 24.0 | 57.0 |
| Body Mass Index [kg/m²] | 23.3 | 3.13 | 17.9 | 31.0 |
| Skin Tone [Fitzpatrick Scale] | 3.10 | 0.74 | 2.00 | 5.00 |
| Earlobe Thickness [mm] | 2.52 | 0.33 | 1.76 | 3.22 |
| Sex | 14 female, 16 male |  |  |  |

**Figure 2:**
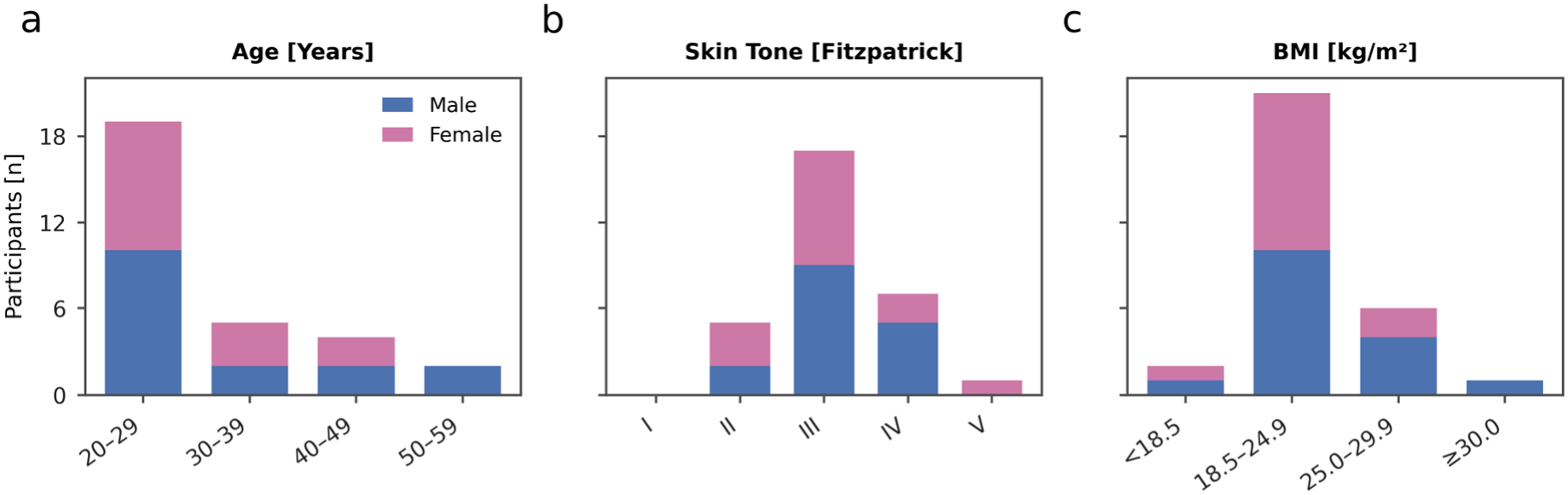
Participant characteristics. (a) Age, (b) skin tone according to the Fitzpatrick scale and (c) body mass index (BMI) of the study cohort. Bars show participant counts, stratified by sex.

### Exploratory Evaluation of the OMS

Each MiROS measurement was temporally aligned such that the signal feature corresponding to the skin surface appeared at the same time point across all measurements. The measurements were further truncated at the time-of-flight corresponding to an apparent depth of 150 µm. We then used this representation as input to a shallow convolutional neural network (CNN) calibrated against the reference blood glucose concentration. The resulting model output was derived retrospectively and was contrasted to the blood glucose values obtained with the glucometer. To minimize bias, model performance was assessed using threefold cross-validation, in which a single model was fitted to two thirds of the pooled experimental data and used to predict the remaining third. In this exploratory analysis, the OMS achieved a MAE of 11.9 mg/dL, a MAPE of 10%, and more than 99.9% of predictions in zones A and B of the Consensus Error Grid (CEG) (Fig. 3a). In this dataset, the OMS’ errors were lower than those of a simultaneously acquired, commercially available CGM, which yielded a MAE of 15.7 mg/dL and a MAPE of 12% (Fig. 3c).

**Figure 3:**
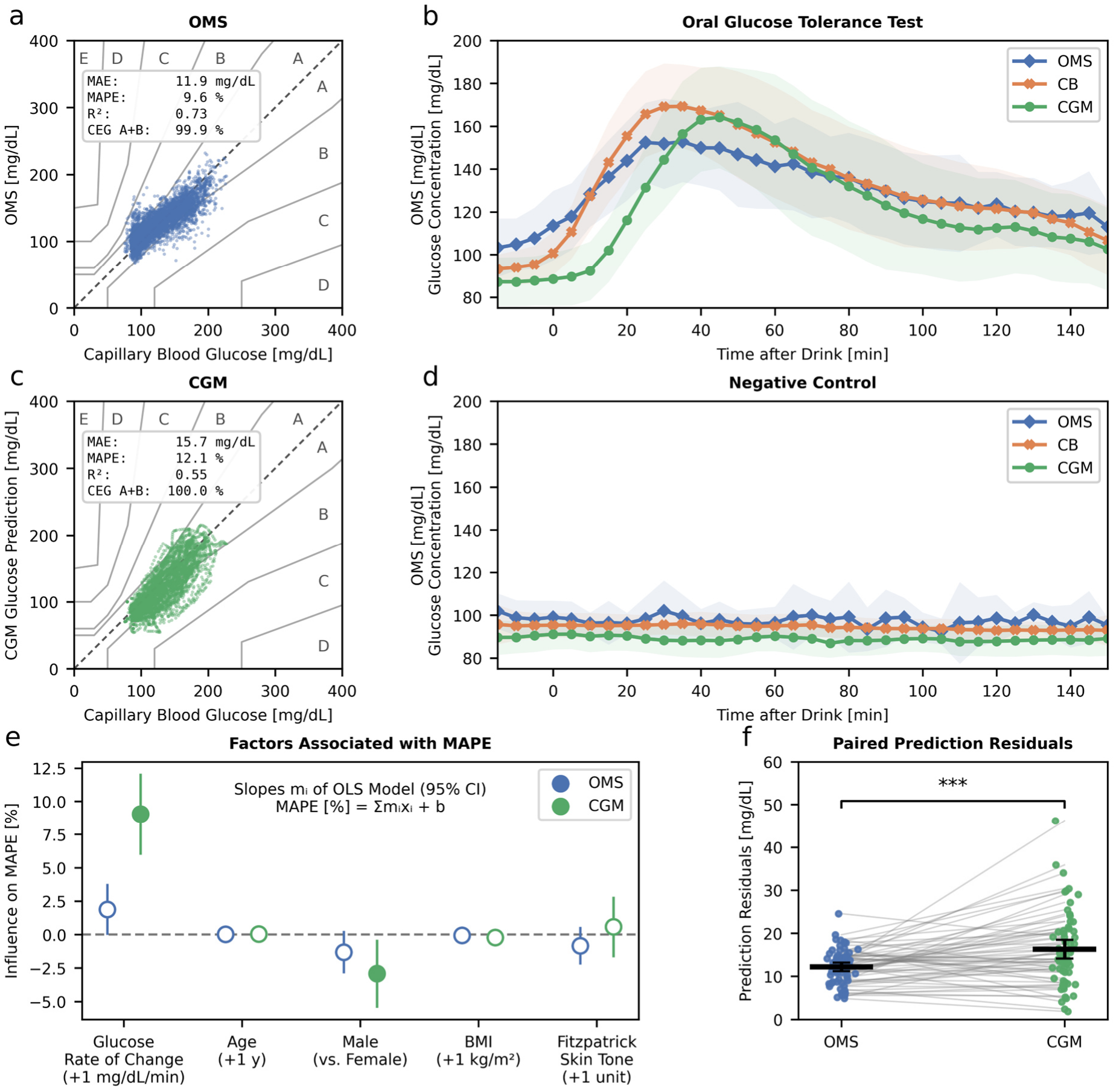
Retrospective evaluation of the Optoacoustic Mid-infrared Signal (OMS). (a) Consensus Error Grid (CEG) for the OMS and capillary blood reference measurements. Dots indicate individual measurement points (n = 5460). (b) Mean and standard deviation of the OMS as well as glucose values obtained in all experimental sessions from the continuous glucose monitor (CGM) and capillary blood reference measurements (CB) after ingestion of glucose solution. (c) CEG for CGM glucose measurements evaluated against capillary blood reference measurements. Dots indicate individual measurement points (n = 5460). (d) Mean and standard deviation of the OMS as well as glucose values obtained in all experimental sessions from the CGM and capillary blood reference measurements after ingestion of water. (e) Coefficients of an ordinary least squares (OLS) model with 95% confidence intervals, fitted to the mean absolute percentage error (MAPE) of each measurement session using participant characteristics as explanatory variables. Filled circles indicate significant effects (p < 0.05). (f) Distribution of prediction residuals for OMS and CGM measurements. (***: p < 0.001, two-sided Wilcoxon test)

Across the pooled cohort, the OMS response related closely to the time course of the capillary blood reference measurements (Fig. 3b). Averaged over all OGTT sessions, the OMS showed a time lag close to zero relative to capillary blood glucose (mean 0.4 min; interquartile range, IQR, −3.8 to 2.5 min), whereas the CGM lagged by 10.5 minutes (IQR 8.8 to 12.5 min), as is typical for ISF glucose. This lack of delay is consistent with our previous study in mice^6^, in which depth-gated MiROS signals reflected tissue changes closely coupled to vascular glucose dynamics.

We also examined whether participant characteristics influenced the relative deviation of the OMS from the blood glucose and the CGM readings. Ordinary least squares modelling indicated that CGM accuracy was associated with both the rate of change of blood glucose concentration (p < 0.001) and participant sex (p = 0.02), with higher error in female participants, whereas no participant characteristic showed a statistically significant association with MiROS relative accuracy (all p > 0.05) (Fig. 3e). Consistent with these findings and previous studies^7^, the OMS showed a significantly lower mean absolute measurement error than the CGM (p < 0.001, two-sided Wilcoxon test; Fig. 3f).

In an additional analysis we asked whether the OMS can be derived without session-specific calibration, using leave-one-session-out cross-validation. Calibration-free derivation was successful (R^2^ > 0) in 12 of the 62 experimental sessions. Within this subset, the OMS achieved an MAE of 15.5 mg/dL and a MAPE of 12%, with 80% of OMS values located in zone A and 100% in zones A and B of the CEG (Fig. 4a). To explore which factors were associated with successful calibration-free derivation of the OMS, we performed logistic regression analysis. Participant characteristics (BMI, sex, age, skin tone) as well as glucose rate of change and glucose excursion span showed no statistically significant association with derivation success (all p > 0.05). In contrast, higher signal-to-noise ratio (SNR) was significantly associated with a greater likelihood of successful calibration-free prediction (p = 0.01, Fig. 4b,c), suggesting that the small glucose-related signal may only be exploited without session-specific calibration when it is not masked by non-glucose temporal fluctuations. These findings identify SNR as an important, but possibly not exclusive, determinant of calibration-free prediction performance.

**Figure 4:**
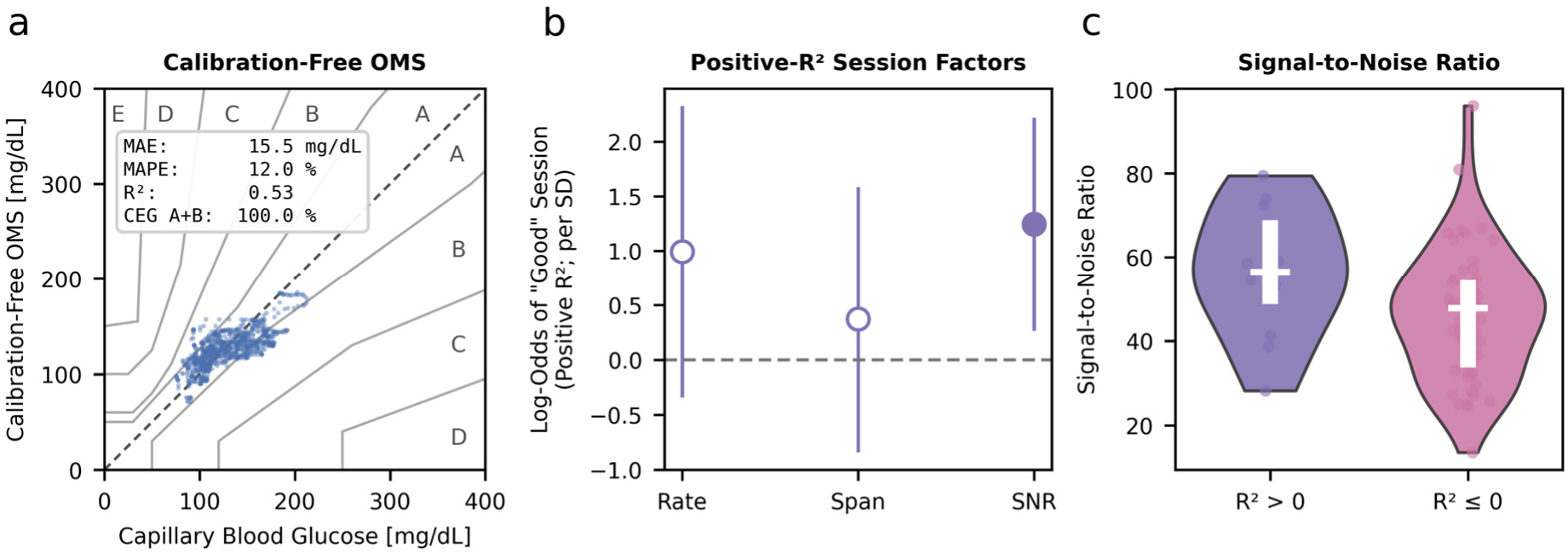
Evaluation of the Optoacoustic Mid-infrared Signal (OMS) without prior calibration and determinants of measurement success. (a) Consensus Error Grid (CEG) for the calibration-free OMS in a reduced subset of 12 experimental sessions, evaluated against capillary blood reference measurements. (b) Log-odds coefficients with 95% confidence intervals from a logistic regression model distinguishing sessions with successful calibration-free OMS derivation from sessions in which calibration-free derivation was not possible. Glucose rate of change, glucose excursion span, and signal-to-noise ratio were included as explanatory variables. (c) Distribution of the signal-to-noise ratio from sessions with positive (R^2^ > 0) and negative (R^2^ < 0) coefficients of determination for calibration-free glucose prediction.

## Discussion

In this study, we demonstrate that MiROS responses correlate well with blood glucose concentrations in an OGTT context in healthy individuals. This finding translates recent evidence from animal experiments into human measurements and supports MiROS as a promising step toward a future non-invasive glucose monitoring approach. In particular, we found that the OMS correlated with capillary blood glucose changes during OGTTs with a MAE of 11.9 mg/dL and a MAPE of 10%. These errors were lower than those of a simultaneously acquired commercial CGM (MAE 15.7 mg/dL, MAPE 12%). In addition, the timing of the OMS signal showed a close relation to the dynamic changes observed in blood glucose by the glucometer measurements, without the characteristic delay of CGM, which measures interstitial glucose values. Averaged over all OGTT sessions, the OMS lag was found to be 0.4 min, compared with 10.5 min for the CGM. These observations are consistent with the physiological delay expected between blood glucose and interstitial glucose^1–4^. The observed error values were generally lower than in our initial demonstration in mice (20-50 mg/dL root mean square error)^6^, despite the fact that a single model was fitted to the pooled data of all participants, whereas the mouse study relied on animal-specific calibration. This may be due to a larger blood volume sampled in human skin compared with mouse skin. Nevertheless, we note that while the OMS was implicitly calibrated against the same general capillary reference used for its evaluation, the CGM is factory calibrated. The comparison with the CGM should therefore be read as a pilot comparison and not as a performance benchmark. Still, reaching this level of agreement and timing in the first human measurements with a MiROS prototype is encouraging and supports prospective evaluation in larger, more diverse cohorts. We further found the OMS accurately mapped onto the blood glucose values without any prior calibration on a subset of 12 of the 62 sessions. Within these sessions the OMS correlated with blood glucose (R^2^ > 0), with an MAE of 15.5 mg/dL and all values in zones A and B of the CEG.

As this study represents the first *in vivo* evaluation of MiROS in humans using a first prototype system, measurement stability and reproducibility were not yet fully optimized. The limited success of calibration-free derivation is likely due to a combination of measurement noise, temporal drift and inter-session variability. These may arise from different factors, including variations in acoustic coupling and tissue acoustic properties, especially since in its current setup, the prototype requires precise spatial confocal alignment of mid-infrared excitation and ultrasound detection over a separation of approximately 4 mm, making the measurement sensitive to small changes in probe position, tissue motion, and acoustic coupling. In addition, the measured signal is a smoothed representation of the actual absorbed energy distribution within the tissue, shaped by the transducer impulse response^8^, the excitation pulse profile^9^ and frequency-dependent attenuation^10^, which mixes signals from glucose-bearing layers with superficial layers of low glucose content. As skin structure differs between participants, this dilution of the glucose-specific signal is participant-specific, limiting the overall accuracy of the measurements. Nevertheless, we recently demonstrated that the underlying method remains stable over periods exceeding one week and is robust to previously unseen spectral backgrounds^11^. These observations point to a future system design that improves the SNR and measurement reproducibility, by improving the geometrical arrangement and advancing excitation and detection hardware^12,13^.

We note that in contrast to recent non-invasive studies using Raman or mid-infrared optothermal spectroscopy in diabetic individuals^14–16^, the present study investigated smaller glucose variations observed during OGTTs in healthy individuals. Blood glucose concentrations in other studies reached approximately twice the maximum values observed here. Instead, the OMS was evaluated across blood glucose concentrations of 75-225 mg/dL, a range relevant to physiological glucose dynamics. Despite these constraints, MiROS achieved comparable accuracy while requiring neither session-specific pre-conditioning^15^ nor prolonged acquisition times^16^. These findings support MiROS potential for non-invasive monitoring under conditions involving comparatively subtle glucose changes, possibly improving upon the performance of other non-invasive approaches, including methods that operate on tears^17^, saliva^18^, or sweat^19^ or optical approaches that also offer measurements primarily in ISF, by using optical polarization^20^, elastic scattering^21^, Raman scattering^14^, or optical absorption spectroscopy in the near infrared^22^, mid-infrared^15^ or terahertz ranges^23^. MiROS capitalizes on the high absorption cross-sections of glucose in the mid-infrared^24^ in combination with the depth-resolving capability of optoacoustic detection. In contrast to purely optical mid-infrared approaches^15^, which integrate the signal over superficial skin layers, this allows MiROS to preferentially sample vascularized layers such as the capillary loop layer at the epidermal-dermal junction.

In summary, in this pilot study, MiROS signals obtained from a human study showed good correlation to blood glucose dynamics without the delay of interstitial glucose measurements as performed by CGM. Sensor designs that minimize motion effects and measurement noise, together with larger and more diverse training data, are next steps that can further improve performance.

## Methods

### MiROS System

The MiROS system was built around an external-cavity quantum cascade laser (Hedgehog, Daylight Solutions, USA) operating over a tunable wavenumber range of 1250-923 cm^-1^ at a pulse repetition frequency of 100 kHz. The excitation beam was focused onto tissue using an aspheric lens. Ultrasound waves generated through the optoacoustic effect were detected in transmission geometry by a focused ultrasound transducer (central frequency, 25 MHz) confocally aligned with the optical focus. Spectral acquisition was performed by sequential wavelength tuning, with 1000 optoacoustic signals averaged per wavelength and 10 spectra subsequently averaged. The transducer signal was digitized at 500 MSPS with a data acquisition card (CSE50216, GaGe, USA). System control, data acquisition, and automated measurement execution were implemented in Python.

### Optoacoustic Mid-Infrared Signal

A shallow convolutional neural network (CNN) was used to relate the OMS to reference glucose concentrations. To reduce scaling variability caused by changes in coupling or temperature, the OMS was normalized by its Euclidean norm. After per-element standardization, model performance was assessed in two evaluation settings: threefold cross-validation, including support data from every session (Fig. 3), and leave-one-session-out cross-validation, simulating a fully calibration-free deployment scenario (Fig. 4). All preprocessing parameters, including per-element standardization coefficients, were estimated exclusively from the training partition and applied unchanged to the corresponding test partition. MAE, MAPE, and R^2^, as well as the fractions of predictions within the respective CEG zones, were computed as the primary outcome measures in Python. The absolute errors of the OMS and the CGM relative to the capillary reference were compared using a two-sided Wilcoxon signed-rank test, with a significance level of α = 0.05.

### Regression Analysis of MAPE and Calibration-Free Model Performance

Statistical analyses were performed using the Python library statsmodels. Ordinary least squares regression was used to investigate the association between MAPE and predefined participant characteristics (Fig. 3e). To identify determinants of calibration-free model performance, we applied logistic regression to assess the influence of glucose excursion metrics, including the span and rate of change of the excursion, as well as system signal stability, on the probability of a session being classified as successful, i.e. a session achieving an R^2^ value exceeding 0 (Fig. 4b). Signal stability was quantified for each session as a temporal SNR. For every element of the depth- and wavenumber-resolved optoacoustic signal, we divided its mean over the session by its standard deviation over the session. We then averaged these ratios across all elements to obtain one SNR value per session.

### Time-Lag Analysis

To quantify the temporal delay relative to capillary blood glucose, we estimated the lag of the CGM and of the OMS separately for each glucose-ingestion session. Within a session, the capillary reference and the test signal were aligned on time after drink ingestion and linearly interpolated onto a uniform time grid with spacing equal to the median sampling interval. The Pearson correlation between the reference and the time-shifted test signal was calculated for all shifts within ±30 min, and the shift with the highest correlation was taken as the lag of that session. The OMS was evaluated on its cross-validated predictions and smoothed with a Savitzky-Golay filter (second-order polynomial, window of 20 samples) before lag estimation. Lags are reported as mean and interquartile range across sessions.

### Study

The measurements were collected in a clinical study (approved by the TUM Ethics Committee and registered with the German Clinical Trials Register DRKS00036371) in accordance with the Declaration of Helsinki. All participants signed an informed consent before voluntary inclusion. The study was conducted as a single-center pilot investigation in healthy participants to evaluate the association between MiROS signals and glucose variations determined by standard reference measurements. Participants were required to be 22-60 years of age, to arrive fasting for at least 12 h, and to refrain from smoking on the day of the measurement. Participants underwent MiROS measurements in parallel with capillary blood glucose measurements and a commercial minimally invasive CGM (Freestyle Libre 3, Abbott Diabetes Care, USA). The CGM had been attached at least 3 days before the first measurement session according to the manufacturer’s instructions. In total, 30 participants were investigated to assess the relationship between MiROS signals and reference blood glucose measurements, and each participant was measured on at least two separate study days. One session consisted of MiROS measurements following ingestion of 75 g glucose equivalent in 300 ml liquid. In the second session, a randomly selected subgroup (approximately one third of participants) received water as a negative control, while the remaining participants received a second glucose solution. Before the start of the first measurement session, participant earlobe thickness was measured using a micrometer screw and participants’ skin tone on the Fitzpatrick scale was determined using a skin tone analyzer (GP531, DEESS, China).

### MiROS Measurement Protocol

On the measurement day, participants were positioned in a comfortable seated or supine posture and rested for at least 15 min before data acquisition. MiROS measurements were performed on the earlobe, which was prepared by cleaning with isopropyl alcohol and tape strips to reduce interference from the skin surface. MiROS measurements were performed quasi-continuously throughout the session and recorded in parallel with repeated capillary blood glucose measurements (Contour Next, Ascensia Diabetes Care GmbH, Germany). Baseline measurements were obtained from -15 min to 0 min in approximately 5-min intervals, after which participants ingested either 75 g glucose equivalent in 300 ml liquid or water at t = 0 min. Capillary blood sampling then continued every 2-5 min up to 150 min.

## Data Availability

All data produced in the present study are available upon reasonable request to the authors

## Abbreviations

BMI: Body Mass Index
CEG: Consensus Error Grid
CGM: Continuous Glucose Monitor
CNN: Convolutional Neural Network
IQR: Interquartile Range
ISF: Interstitial Fluid
MAE: Mean Absolute Error
MAPE: Mean Absolute Percentage Error
MiROS: Mid-Infrared Optoacoustic Sensing
OLS: Ordinary Least Squares
OMS: Optoacoustic Mid-infrared Signal
OGTT: Oral Glucose Tolerance Test
QCL: Quantum Cascade Laser
SNR: Signal-to-Noise Ratio

## Acknowledgments

This project has received funding from the European Union’s Horizon 2020 research and innovation programme under grant agreement No 862811 (RSENSE) and from the European Union’s Horizon Europe research and innovation programme under grant agreement No 101058111 (GLUMON).

